# Pathway Modeling of Genomic and Tissue-Specific Transcriptomic Architecture Identifies Personalized Mechanisms of Atrial Fibrillation Risk^†^

**DOI:** 10.64898/2026.08.25.26361369

**Authors:** Rasika Venkatesh, Rajat Deo, Thomas P. Cappola, Penn Medicine BioBank, Marylyn D. Ritchie, Dokyoon Kim

## Abstract

Atrial fibrillation (AF) is the most common sustained cardiac arrhythmia and a major cause of cardioembolic stroke. Although polygenic risk scores (PRS) are well characterized to quantify inherited susceptibility for AF, they provide limited insight into the pathways and tissues underlying genetic risk, which are critical to uncover for individual risk prediction. In this study, we develop a pathway-level multi-omics representation learning framework that converts individual genetic profiles into interpretable biological features by integrating GWAS-derived pathway burden scores with tissue-specific transcriptomic pathway signals. We constructed machine learning models to assess population-level AF risk prediction performance across genomic and transcriptomic tissue contexts; the pathway-based global attention models substantially improved risk prediction performance over PRS and other baselines (AUROC improved from 0.601 to 0.738). Transformer and graph neural network frameworks then assessed individual-level pathway interpretability, revealing heterogeneous contributions from electrical signaling, cardiac development, and DNA repair pathways to AF risk. This added interpretability highlights the potential of this pathway approach to enable more mechanistically informed risk stratification than static PRS by capturing underlying heterogeneity. To independently assess whether prioritized pathways reflected cardiac regulatory biology, we compared pathway rankings with transcriptional effects predicted by the AlphaGenome foundation model. Variants in highly ranked pathways showed significantly greater predicted effects on expression in atrial and ventricular tissues (FDR = 0.032) relative to controls, providing orthogonal evidence that the model identifies biologically relevant mechanisms. Overall, this work reframes polygenic risk from a single measure of susceptibility to tissue-informed pathway mechanisms, providing a framework for interpretable genomic stratification in complex diseases.

## 1. Introduction

### 1.1. Atrial Fibrillation (AF) presents high clinical burden despite genetic advances

Atrial fibrillation (AF) is the most common sustained cardiac arrhythmia, affecting over 37 million individuals worldwide, with a lifetime risk approaching 1 in 3 adults^1,2^. AF substantially increases an individual’s risk of heart failure, ischemic stroke, cognitive decline, and premature mortality, resulting in significant clinical burden^2^. Despite advances in rhythm control strategies, monitoring, catheter ablation, and anticoagulation, current approaches for identifying AF largely depend on evidence of structural and electrical remodeling^2^. As a result, there are limited opportunities for early intervention. Large-scale genome-wide association studies (GWAS) have identified over 100-140 susceptibility loci, highlighting the highly polygenic nature of AF^3^. Key loci include rs2200733 and rs10033464 near *PITX2* on chromosome 4q25, rs13376333 in *KCNN3* on 1q21, and rs7193343 in *ZFHX3* on 16q22, which have been replicated across populations^3–5^. These loci map to diverse biological processes, including cardiac electrophysiology (*KCNN3, KCND3*), structural remodeling (*TTN*), inflammation, and cardiac development (*PITX2, NKX2-5*)^6–8^. However, individual variants confer modest effect sizes, and a substantial proportion of heritability remains unexplained^9^. Functional interpretation of GWAS signals is limited by context-specific gene regulation and incomplete understanding of variant effects in atrial tissues^10^. As such, translating genetic associations into biological mechanisms remains a central challenge.

### 1.2. Pathway-based representations enable biologically interpretable risk prediction

Genetic risk scores and polygenic risk scores (PRS) aggregate the effects of known disease-associated variants into static measures of genetic predisposition towards disease. These scores can stratify individuals at elevated risk of developing AF prior to onset^11,12^. One study developed a PRS based on 142 independent AF-associated variants which showed a 54% higher risk per standard deviation^13^. In patients with cardiovascular disease, AF PRS improved the C-index from 0.65 (clinical risk score alone) to 0.70 with PRS addition^14^. However, conventional PRS models demonstrate substantial heterogeneity at the individual level, and individuals with the same risk scores may harbor different underlying mechanisms^15^. The lack of mechanistic interpretability means that even a high PRS provides little actionable information regarding underlying biological processes driving susceptibility. Consequently, while PRS may identify population subgroups warranting enhanced monitoring, it remains inadequate for individualized risk prediction^15^.

Biological pathways provide an intermediate representation between individual variants and disease phenotypes by aggregating distributed genetic effects into functional processes. AF-associated genes have been enriched in cardiac conduction within His-Purkinje cells, action potential generation and repolarization, potassium channel activity, actin-mediated cell contraction, among others^10,16,17^. Co-expression analyses in human atrial tissue further implicated oxidative stress and cell signaling as key pathways, with candidate AF genes overrepresented in modules related to cell injury, developmental processes, metabolic function, and immune activation/inflammation^18–20^. Pathway-based PRS approaches extend this idea by testing whether polygenic burden is enriched in biologically annotated gene sets, offering a more interpretable framework than variant-level scoring alone^21^. However, these methods also remain dependent on pathway definition and annotation quality, and they may improve interpretability more than they improve absolute individual-level prediction.

### 1.3. Representation learning strategies enable biological discovery beyond prediction

Recent advances in deep learning, particularly representation-learning models applied to high-dimensional biological and clinical data, have demonstrated strong performance across prediction tasks by capturing complex nonlinear dependencies that traditional statistical models may overlook. One study found that ECG-AI-derived AF risk reflected inherited susceptibility, with signals at established AF loci. The model predicted incident AF with a hazard ratio 1.06, outperforming baseline PRS^22^. However, such models are often limited in interpretability, raising concerns about whether their predictions reflect real biological mechanisms^22^. Emerging approaches utilizing attention mechanisms and feature attribution provide opportunities to reconcile predictive performance with biological interpretability.

In this study, we developed a pathway-based modeling framework integrating graph neural networks, transformer architectures, and global attention mechanisms to predict atrial fibrillation risk. Our approach incorporates GWAS-derived pathway scores, tissue-specific transcriptomic pathway scores, and PRS to capture complementary dimensions of genetic risk. We evaluated whether transcriptomic augmentation improves predictive performance and, critically, whether these models identify biologically coherent mechanisms at both population and individual levels. To enhance robustness and biological validity, we combined multiple interpretability strategies and performed independent validation using the AlphaGenome foundation model^23^. Using this integrative approach, we aimed to determine whether genetically informed pathway-level representations can improve mechanistic understanding of AF while maintaining or enhancing predictive accuracy, thereby advancing the translational potential of genetic risk modeling.

## 2. Methods

### 2.1. Penn Medicine BioBank AF Study Population

We conducted all analyses in the Penn Medicine BioBank (PMBB), a health system-linked biobank of 57,170 genotyped participants with longitudinal electronic health record (EHR) data^24^. Genotypes were imputed using the TOPMed reference panel (Version r2, 2020), and participants provided informed consent for genomic analyses. Genetic ancestry was inferred via principal component analysis (PCA) projection onto 1000Genomes Phase 3 reference populations^25,26^. Demographic covariates used in risk prediction were selected based on their association with AF status (**Table 1)**. AF cases were defined as individuals with at least 2 AF diagnostic codes (ICD10 427.2, 427.3*) recorded during separate clinical encounters^27^. Controls were defined as individuals with no AF codes; individuals with a single code were excluded as phenotypically ambiguous. Age was defined as age at first AF diagnosis for cases, and age at most recent EHR encounter for controls. The final cohort comprised 52,440 individuals (N=10,585 cases; 20.4% prevalence).

**Table 1.** Demographic and clinical information for the study population. P-values calculated using Pearson’s Chi-Squared Test (Sex, Ancestry) and Welch’s T-Test (Age)

| Variable | Cases (n = 10,694) | Controls (n = 41,746) | P-Value |
| --- | --- | --- | --- |
| **Sex** |  |  | <0.001 |
| Male | 7,388 (69.1%) | 19,224 (46.0%) |  |
| Female | 3,306 (30.9%) | 22,522 (54.0%) |  |
| **Ancestry** |  |  | <0.001 |
| African (AFR) | 1,682 (15.7%) | 10,178 (24.4%) |  |
| Admixed American (AMR) | 66 (0.2%) | 640 (1.5%) |  |
| European (EUR) | 8,823 (82.5%) | 29,692 (71.1%) |  |
| East Asian (EAS) | 63 (0.6%) | 594 (1.4%) |  |
| South Asian (SAS) | 60 (0.6%) | 642 (1.5%) |  |
| **Age** | 64.4 ± 12.5 | 57.7 ± 16.6 | <0.001 |

### 2.2. PMBB GWAS and PRS Calculation

Multi-ancestry and ancestry-stratified (EUR, AFR) genome-wide association studies (GWAS) for AF within PMBB were performed using Saige v1.4.5^28^. Logistic regression with Firth correction accounted for case-control imbalance, with covariates including age at enrollment, sex, and PCs 1-5. Variants with MAF <0.005 within each ancestry were excluded. Genomic inflation (λ_GC_) was assessed; the observed values of 0.85 - 0.98 are consistent with conservative Firth-corrected test statistics^28^. Pre-computed AF PRS weights from a large-scale meta-analysis from the PGS Catalog were applied to PMBB genotypes using PLINK --score, and the resulting score was standardized within the training data^3,29^. The PRS was evaluated with 3 logistic regression models, unregularized, LASSO (L1), and Elastic Net (EN), after splitting the dataset into 70:20:10 train, test, and validation sets balanced by case/control ratio^30,31^. Performance was evaluated using the area under the receiving operator characteristic (AUROC), precision-recall curve (AUPRC), balanced accuracy (BA), and F1-score with 95% confidence intervals estimated from 1,000 bootstrap resamples.

### 2.3. Pathway Feature Construction

#### 2.3.1. GWAS pathway burden scores

Independent lead SNPs from multi-ancestry GWAS were identified by LD clumping (P < 5E-06, R^2^ < 0.1, 500-kb window); 76 of which were mapped to 458 protein-coding genes (±500 kb, GENCODE v38)^32^. They were assigned to pathways from GO Biological Processes, Reactome, and KEGG Legacy datasets of MSigDB v2026.1, yielding 1,654 pathways with 3–30 independent GWAS-implicated genes each^33–38^. For pathway *k*, the GWAS burden score for individual *i* was calculated as a pathway size-normalized, significance-weighted sum of lead-SNP dosages:

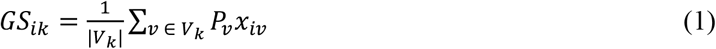

where *x*_*iv*_ is genotype dosage (0–2 copies of effect allele), *p*_*v*_ is the -log(P) of the GWAS signal per variant, and *V*_*k*_ is the set of LD-independent lead SNPs mapped to genes within pathway *k*. Scores were standardized (mean 0, unit variance) on the training set before model fitting.

#### 2.3.2. Tissue-specific transcriptional pathway scores

Genetically regulated expression (GReX) was estimated via PrediXcan using PredictDB models trained on GTEx v8, generating predicted expression for five tissues: heart atrial appendage (HAA), heart left ventricle (HLV), artery aorta (AA), artery coronary (AC), and whole blood (WB)^39,40^. Gene expression values per tissue were aggregated to pathway activity scores:

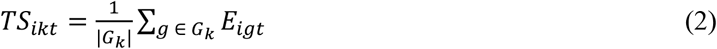

where *E*_*igt*_ is the predicted expression of gene *g* for individual *i* in tissue *t*. Single-tissue models were run for all five tissues; based on sparsity of the L1 and EN models, combined multi-tissue feature sets (all tissues; HAA + HLV; and HAA + HLV + AA) were evaluated.

### 2.4. Interpretable Modeling

#### 2.4.1. Model Architectures and Feature Sets

We evaluated 7 unique model types: logistic regression (LR; unregularized, L1, and EN), random forest (RF), a global attention model, a transformer, and a graph neural network (GNN) to assess the value of pathway features for binary AF risk prediction and downstream interpretability. Each participant was represented as a *K×T* pathway matrix (1,654 pathways across *T* feature layers)

(**Table 2**). Clinical covariates (age, sex, ancestry PCs 1–5) and the PRS were appended as flat covariates rather than pathway features. Before model fitting, variance-based feature-selection retained the top 500 pathways by training-set variance. Variance was rank-normalized within each feature type when multiple feature types were combined, so that GWAS and GReX score variances, which differ by orders of magnitude, did not asymmetrically dominate the ranking. This filter was applied uniformly to every model architecture and every pathway-containing feature set.

**Table 2.** Model feature sets and total number of features.

| Feature Sets | Total Number of Features | Models |
| --- | --- | --- |
| PRS | 1 | LR (unregularized, L1, EN) |
| PRS + Covs | 8 | LR (unregularized, L1, EN) |
| Covariates (Age, Sex, PCs) | 7 | LR, RF, Global Attention, Transformer |
| GWAS pathway scores + Covs | 1654 + 7 (500 pathways) | LR, RF, Global Attention, Transformer |
| GWAS pathway scores + PRS + Covs | 1654 + 8 (500 pathways) | LR, RF, Global Attention, Transformer |
| GReX pathway scores + Covs (each tissue) | 1654 + 7 (500 pathways) | LR, RF, Global Attention, Transformer |
| GReX pathway scores (all tissues) + Covs | (1654 * 5) + 7 (500 pathways) | LR, RF, Global Attention, Transformer |
| GWAS + GReX (HAA, HLV) + Covs | (1654 * 3) + 7 (500 pathways) | LR, RF, Global Attention, Transformer, GNN |
| GWAS + GReX (HAA, HLV) + PRS + Covs | (1654 * 3) + 8 (500 pathways) | LR, RF, Global Attention, Transformer, GNN |
| GWAS + GReX (HAA, HLV, AA) + Covs | (1654 * 4) + 7 (500 pathways) | LR, RF, Global Attention, Transformer, GNN |
| GWAS + GReX (HAA, HLV, AA) + PRS + Covs | (1654 * 4) + 8 (500 pathways) | LR, RF, Global Attention, Transformer, GNN |

Participants were split using stratified random sampling by AF status into training, validation, and test cohorts (70:10:20), maintaining a case prevalence of 20.4% in each set^41^. Hyperparameter optimization was performed only on training and validation data, with final evaluation on the held-out test set. Model discrimination was assessed using AUROC, AUPRC, Brier score, F1 score, and calibration curves, with 95% confidence intervals estimated from 1,000 bootstrap resamples.

#### 2.4.2. Logistic Regression Models (Unregularized, LASSO, Elastic Net)

Three logistic regression models, unregularized, L1, and EN, were fitted using the SAGA optimizer with class-weight balancing, and continuous predictors were standardized using training-set statistics^41^. Regularization strength was selected by five-fold cross-validation using validation AUROC over a logarithmically spaced grid of C values (C = {1E-04 - 1E-02}). The EN model additionally optimized the L1/L2 mixing parameter (L1 ratio = {0.1, 0.5, 0.7, 0.9})^31^.

#### 2.4.3. Random Forest

A 500-tree random forest served as the nonlinear ensemble baseline. Maximum tree depth ({8, 12, 15, None}) and minimum samples per leaf ({20, 50}) were selected by grid search against held-out validation AUROC (a single train/validation evaluation per candidate, not k-fold CV); the number of features considered per split used the scikit-learn default^41,42^. Feature importance was quantified by Gini index, aggregated to pathways for comparison with attention-based rankings^43^.

#### 2.4.4. Global Pathway Attention Model

The global pathway attention model was designed to learn a single population-level pathway importance vector. Each pathway was independently embedded using a shared encoder consisting of 2 Linear–LayerNorm–ReLU–Dropout blocks (embedding dimension = 128). A learned attention vector for each pathway was softmax-normalized to generate pathway weights that were shared across all individuals, producing a weighted sum of pathway embeddings^44–46^. This representation was layer-normalized, concatenated with a 2-layer multilayer perceptron (MLP) embedding of clinical covariates, and passed to a linear classifier. Because the attention weights are model parameters rather than post hoc attribution scores, they provide an intrinsic population-level ranking of pathway importance^45^. Models were trained using AdamW (learning rate = 1E-03, weight decay = 1E-04) with cosine annealing learning-rate scheduling, gradient clipping (maximum norm = 1.0), focal loss (*γ* = 2), and balanced mini-batches generated with a WeightedRandomSampler^47–49^. Embedding dimension and dropout were fixed across feature sets to enable direct comparison of pathway importance.

#### 2.4.5. Transformer

The pathway transformer modeled individualized pathway interactions through self-attention. Pathway embeddings were combined with learned pathway identity embedding; positional encodings were omitted since pathways have no intrinsic ordering^45^. A learnable classification (CLS) token was prepended to the pathway sequence and processed by 2 pre-normalized transformer encoder layers with 4 attention heads (embedding dimension 128, feed-forward dimension 128, dropout 0.2)^45,50^. The final CLS representation was concatenated with an MLP-derived covariate embedding and classified using a linear layer. Training followed the same optimization as the global attention model, with early stopping based on validation AUROC (patience = 15 epochs; maximum = 200 epochs). Predicted probabilities were calibrated using Platt scaling fitted on the validation set before final evaluation on the held-out test set^51^.

#### 2.4.6. Graph Neural Network (GNN)

GNNs were implemented using a 2-layer GraphSAGE architecture to model pathway–pathway relationships^52^. The top 500 pathways were represented as nodes, and 2 graph construction strategies were evaluated: 1) a Jaccard similarity graph connecting pathways sharing at least three genes with Jaccard similarity ≥ 0.1, and 2) a training-set-only k-nearest-neighbor graph (k = 50) based on absolute Pearson correlations between pathway scores. Each GraphSAGE layer incorporated residual connections, LayerNorm, ReLU activation, and dropout (embedding dimension = 128). Global mean and max pooled graph embeddings were concatenated with a covariate embedding and passed to a linear classifier. Training followed the same procedure as the global attention model with a reduced batch size of 32. Individual pathway importance was quantified using GradCAM scores, computed as the ReLU of the gradient–activation product on the final node embeddings and averaged across embedding dimensions^53^. Final predictions were calibrated using Platt scaling.

### 2.5. Biological Interpretation

#### 2.5.1. Population-level Interpretation

Population-level pathway importance was characterized using the global attention model and transformer. For the global attention model, pathway importance was obtained directly from the learned softmax-normalized attention vector. For the transformer, per-individual attention vectors were z-score normalized before averaging across the test cohort^54^. Differential pathway attention between AF cases and controls was assessed using two-sided Mann–Whitney U tests with Benjamini–Hochberg false discovery rate (FDR) correction. Individuals were additionally stratified into deciles according to predicted AF probability, and mean pathway attention was summarized across risk groups for the most strongly case-enriched pathways.

#### 2.5.2. Individual-level Interpretation

Individualized mechanisms were investigated using the transformer model and GNN, both of which generate patient-specific pathway importance profiles. For the transformer, per-individual attention vectors were projected into two dimensions using Uniform Manifold Approximation and Projection (UMAP) to visualize heterogeneity in learned biological representations^55^. In the GNN, pathway importance was quantified using GradCAM attribution, computed as the gradient–activation product on the final GraphSAGE node embeddings and averaged across embedding dimensions^56^. Population-level pathway rankings were obtained by averaging GradCAM scores across the test cohort, while individual attribution profiles were compared across predicted-risk strata^45,56^. Attribution consistency was additionally evaluated between the Jaccard similarity and pathway score-correlation graph construction strategies to distinguish robust biological signals from graph topology–specific effects.

### 2.6. AlphaGenome Foundation Model–Based Biological Validation

Pathway importance scores from the global attention model (GWAS + PRS feature set) were used to rank all scored pathways, with the top 50 and bottom 50 pathways by attention weight selected for AlphaGenome (v0.7.0; Google DeepMind) analysis^23^. GWAS variants were mapped to pathways using a precomputed variant-pathway map derived from 76 LD-clumped AF GWAS lead SNPs. For pathways with multiple variants, variants were prioritized by absolute GWAS effect size (|Beta|). For each variant, AlphaGenome was queried using a 1,048,576 bp sequence window centered on the variant position. Predicted RNA-seq delta scores, representing the difference in predicted gene expression between reference and alternate alleles, were extracted for the 5 GTEx tissues. Variant-level regulatory scores were summarized as mean absolute delta scores and aggregated to the pathway level by averaging across variants assigned to each pathway. Importantly, this analysis was restricted to the GWAS + PRS global attention rankings, which contains no GReX-derived features, to perform an independent assessment of tissue specificity without biasing the model with GReX tissues overweighting themselves. Pathway scores were then compared between the top and bottom pathway tiers using one-sided Mann-Whitney U tests, with FDR correction applied across the 5 tissue tests.

## 3. Results

### 3.1. GWAS Discovery and Feature Construction

The AF discovery GWAS identified 417 genome-wide significant SNPs (p < 5E-08), with the strongest signal on chromosome 4 (p = 2.30E-10), consistent with the 4q25 locus near *PITX2*, the most replicated AF susceptibility locus in the prior GWAS^3^. Significant known loci were also implicated in chromosomes 1, 6, 10, and 16 (**Supplemental Figure S1A**)^3^. Ancestry-stratified analyses (**Supplemental Figure S1B, S1C**) produced similar results in the EUR cohort with decreased significance in the AFR cohort, due to reduced statistical power. Top independent SNPs from GWAS results were subsequently aggregated into pathway-level features for downstream modeling (**Supplemental Table S1**). The highest-ranked pathway scores comprised cardiac-relevant pathways (e.g. atrioventricular valve development, heart valve development, cardiac atrium development, heart morphogenesis, regulation of blood pressure), and broad developmental pathways (e.g. gland, immune system, and nephron development) **(Supplemental Figure S2)**. Tissue-specific GReX features captured orthogonal and complementary information across AA, HLV, AA, AC, and WB tissues **(Supplemental Figure S3)**. Across tissues, 87 - 94% of the highest-ranked pathways were tissue-specific, despite physiological similarity between the cardiac chambers. Well characterized, large pathways such as Gap Junction Assembly and Cell Communication by Electrical Coupling, were consistently prioritized in both cardiac chambers but not in non-cardiac tissues. HAA and HLV shared these 2 highly ranked pathways, while AA and WB tissues exhibited largely distinct pathway profiles. This mixture of disease-relevant and general biological processes motivated model-based interpretability analyses to distinguish AF-specific mechanisms from broad biological pathways.

### 3.2. Predictive Modeling of AF from Pathway Representations

Models were evaluated using combinations of PRS, clinical covariates, GWAS-derived pathway scores, and tissue-specific GReX pathway scores (**Figure 1; Supplemental Table S2)**. The neural architectures (global attention model, transformer, and GNN) consistently outperformed the linear models and RF, with the global attention model and GNN achieving the strongest performance. PRS alone showed limited discrimination (AUROC = 0.601), whereas clinical covariates improved performance (AUROC = 0.697). The largest improvement was obtained by incorporating GWAS pathway scores with PRS, yielding AUROC values of 0.725–0.738 across the models. Similar trends in AUPRC, Brier score, and BA were observed **(Figure 1; Supplemental Figure S4, S5, S6**).

**Figure 1.**
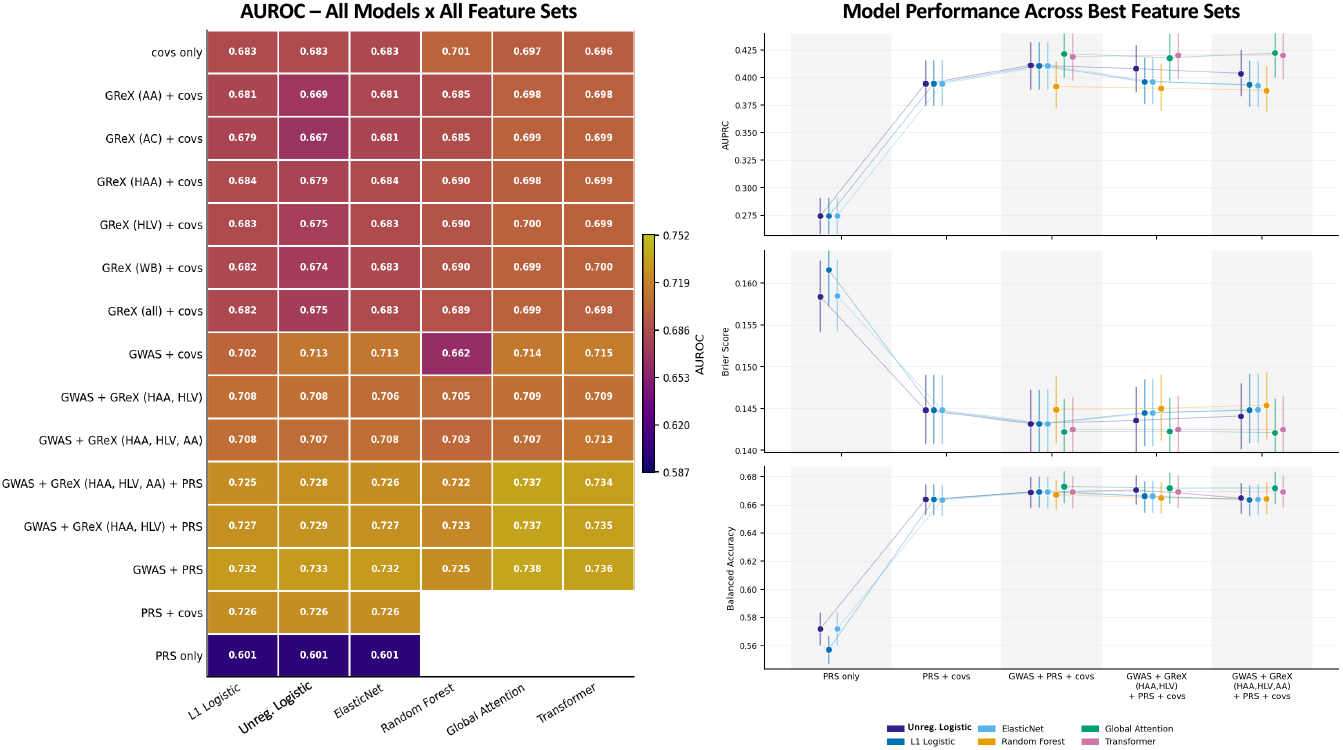
Model performance across all feature sets for L1, L2, EN, RF, global attention, and transformer models. A) Heatmap depicts AUROC values for all models across all feature sets. B) AUPRC, Brier Score, BA depicted in forest plots for representative feature sets (PRS only, PRS + covs, GWAS + PRS + covs, GWAS + GTEx (HAA, HLV) + PRS + covs, GTEx (HAA, HLV, AA) + PRS + covs).

Transcriptomic pathway inclusion maintained predictive performance while providing additional tissue-specific biological information for downstream interpretation. For the global attention model, the GWAS + PRS + covs feature set achieved an AUROC=0.738, AUPRC = 0.421, and Brier score = 0.142. Incorporating HAA + HLV GReX produced similar performance (AUROC = 0.737, AUPRC = 0.419, Brier score = 0.142), as did inclusion of AA (AUROC = 0.737, AUPRC = 0.408, Brier score = 0.142). Due to the 20.4% prevalence of AF in PMBB, absolute AUPRC values remained modest across all models, but represented nearly a 2-fold improvement over the prevalence baseline of 0.204. Since predictive performance was statistically indistinguishable across GReX feature sets, subsequent analyses focused on the HAA + HLV feature set to prioritize model parsimony and cardiac tissue relevance.

The transformer and GNN models achieved comparable predictive performance. The GNN demonstrated similar predictive stability across Jaccard and score correlation graph construction strategies and feature sets (**Supplemental Table S3**). For GWAS + GReX (HAA, HLV) + PRS + covs, the Jaccard topology achieved an AUROC of 0.738, AUPRC of 0.419, and Brier score of 0.143, compared with an AUROC of 0.734, AUPRC of 0.417, and Brier score of 0.142 for the score-correlation topology. Further biological analyses evaluated whether transcriptomic augmentation improves model interpretability while maintaining predictive performance.

### 3.3. Population-Level Biological Interpretation

#### 3.3.1. DNA Repair and Remodeling Mechanisms Characterize Global AF Risk

Population-level interpretation identified biological mechanisms associated with AF risk across the cohort, demonstrating the value of transcriptomic information beyond discrimination. The global attention model provided the primary population-level interpretation because its softmax attention vector is a single model parameter shared across individuals. The transformer pathway rankings were derived by averaging individual attention profiles to provide a complementary view.

The best-performing GWAS + GReX (HAA, HLV) + PRS + covs feature set prioritized pathways in DNA-repair and genome-maintenance machinery, including HDR through Homologous Recombination, Homologous Recombination, Regulation of DNA Repair, and Nonsense-Mediated Decay, alongside metabolic-remodeling pathways such as Pyruvate and Fatty Acid Metabolic Process (**Figure 2A**). Many of these pathways were independently prioritized in the tissue-specific GReX models, supporting their consistency across cardiac transcriptomic contexts. HDR through Homologous Recombination reached the 83–87 percentile in three of five tissues, and most of the remaining DNA-repair pathways and Fatty Acid Metabolic Process appeared in four or five of the five tissue-specific rankings (**Figure 2B**). Covariate attribution identified age (0.150), sex (0.134), and standardized PRS (0.118) as the dominant non-pathway predictors, with ancestry PC1 (0.061) contributing more strongly than PCs 2–5 (0.02–0.05) in the global attention model.

**Figure 2.**
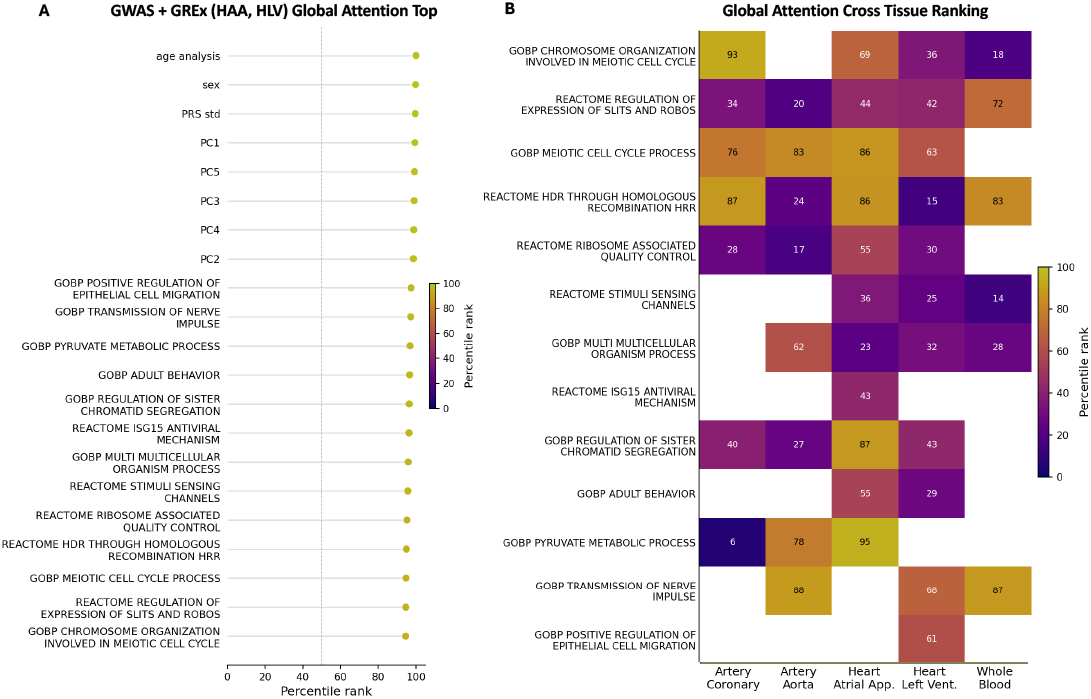
Population-level pathway importance A) Top features identified from global attention GWAS + GReX (HAA, HLV) + PRS + covs. B) Global attention model pathway rankings across all tissues.

Transformer case-control differential attention identified complementary remodeling signals (**Supplemental Figure S7**). The strongest case-enriched pathway under the primary feature set was Sulfur Compound Biosynthetic Process, followed by Negative Regulation of Interleukin-1β Production, linking AF risk to the sulfur compound/H_2_S axis, which has established roles in potassium-channel regulation and suppression of TGF-β-mediated atrial fibrosis. The strongest control pathway was Axon Extension. In the sensitivity analysis incorporating AA tissue, TGF-β Signaling became the dominant case-enriched pathway along with multiple cardiac developmental pathways, suggesting complementary remodeling mechanisms.

#### 3.3.2. Model-specific Attribution Patterns

Transcriptomic augmentation altered attribution behavior in both attention- and graph-based models. Under GWAS + PRS, transformer attention was dominated by Signaling by Moderate Kinase Activity BRAF Mutants, which accounted for approximately 0.25 mean attention (z = 22), consistent with an attention sink. Following incorporation of HAA and HLV GReX, this pattern disappeared; the highest-ranked pathway (Sulfur Compound Biosynthetic Process) reached only z = 3.2, with attention distributed across multiple pathway families. The transformer’s 4 attention heads also showed differentiated behavior; Heads 1–3 demonstrated positive correlations (ρ = 0.25–0.50) and relatively diffuse attention profiles, whereas Head 4 showed negative correlations with the remaining heads (ρ = -0.10 to -0.20) and accounted for the majority of pathway-level differentiation, suggesting specialization of attention heads.

Graph-based interpretation also showed improvement in interpretability. Community detection using the Louvain clustering indicated that the GNN was organized into 43 biologically coherent modules. (**Figure 3A**) These modules included an electrophysiological and structural module centered on Gap Junction Assembly and Cell Communication by Electrical Coupling, as well as developmental signaling (e.g. Dorsal-Ventral Neural Tube Patterning) and DNA repair process modules (e.g. Nucleotide Excision Repair). The full list of modules, labeled by their top pathway, are available in **Supplemental Table S4**. Under GWAS + PRS, GradCAM importance was negatively correlated with pathway node degree (r= −0.29), indicating preferential attribution toward lower-degree independent pathways rather than network hubs. With GReX (HAA, HLV) inclusion, this relationship decreased (r = −0.03), indicating that pathway importance became less dependent on graph topology and less biased toward peripheral pathways **(Figure 3B**). These findings suggest that transcriptomic data reduced attribution artifacts in both transformer and GNN architectures while maintaining performance.

**Figure 3.**
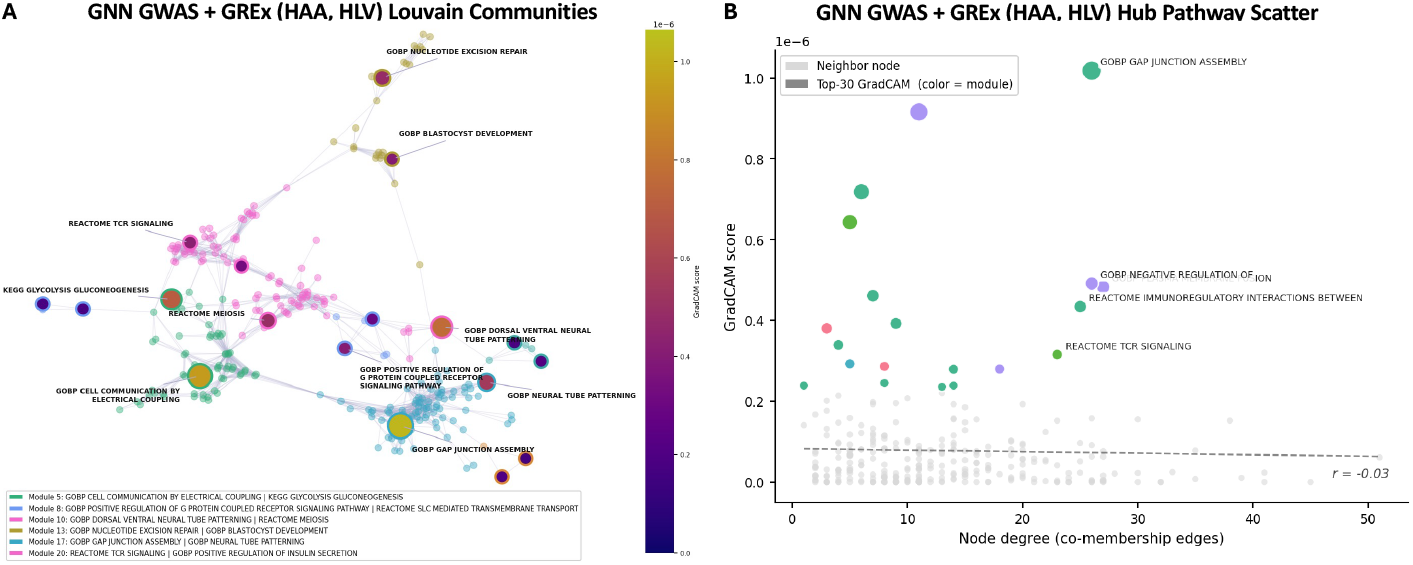
GNN Interpretability. A) Class stratification across PRS tertiles. B) SHAP multiclass feature importance. C) Individual level interpretability

### 3.4. Individual-Level Biological Interpretation

To characterize patient-specific mechanisms underlying AF risk, individual pathway representations were examined to determine whether the model captured heterogeneous biological programs across patients. The embedding organized individuals by pathway usage and predicted risk, with the GWAS + GReX (HAA, HLV) + PRS feature set forming a non-spherical, multi-lobed structure and localized regions enriched for higher-risk individuals (**Figure 4A**). Rather than separating cases from controls, the localized concentrations of higher-risk individuals within specific regions of the embedding indicates that pathway-attention profiles organize patients into risk-associated subgroups rather than along a case/control axis. Risk-decile analysis indicated that pathway contributions varied continuously across predicted risk rather than reflecting only case-control differences. Individuals were ranked into predicted-risk deciles (1,058–1,059 individuals each), and mean pathway attention was compared. Under GWAS + GReX (HAA, HLV) + PRS, Sulfur Compound Biosynthetic Process showed the strongest risk-associated gradient, increasing progressively from D1 to D10 (Δ = 1.65 in z-scored attention), indicating that transcriptomic feature sets emphasize complementary remodeling pathways while preserving patient-level pathway responses (**Figure 4B**).

**Figure 4.**
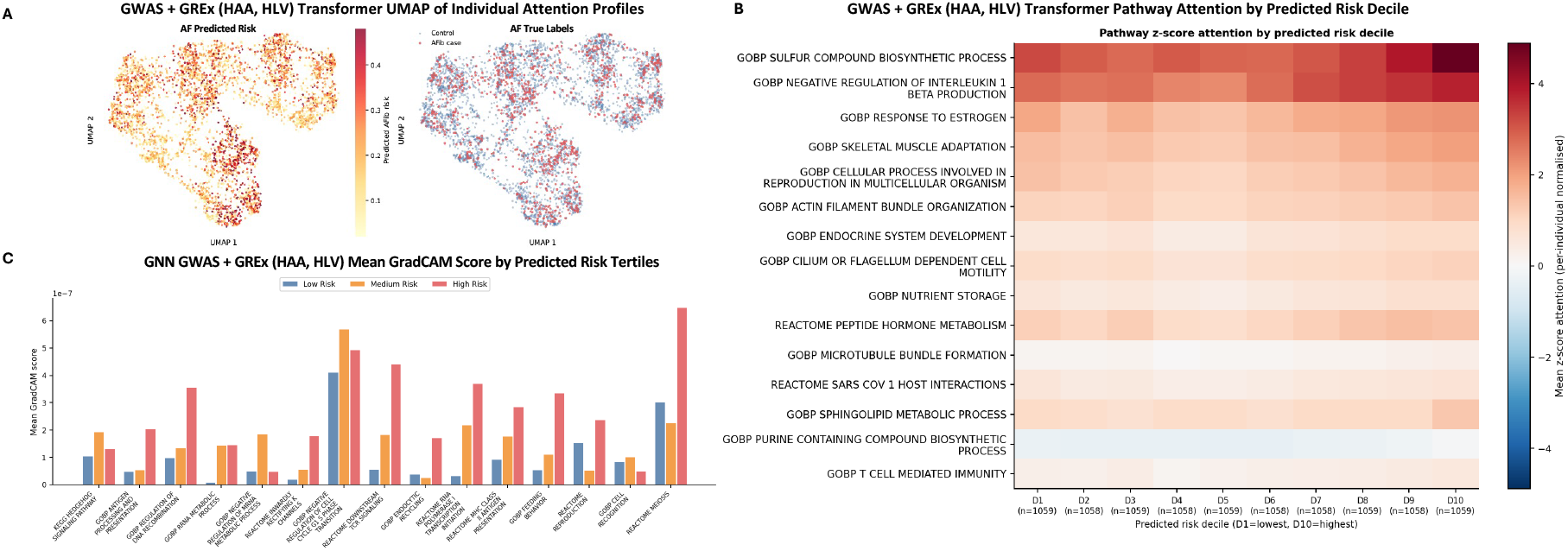
Individual-level interpretability. A) Transformer UMAP of individual predicted risk profiles. B) Transformer pathway-based risk deciles. C) GNN pathway importance by predicted risk tertile

Individual-level pathway importance was also evaluated using GNN GradCAM attribution across predicted-risk tertiles (**Figure 4C**). Under GWAS + GReX (HAA, HLV) + PRS + covs, Inwardly Rectifying K+ Channels increased monotonically from the low- to high-risk tertile, while Endocytic Recycling and Homologous Chromosome Pairing at Meiosis increased between the medium- and high-risk tertiles. Negative Regulation of mRNA Metabolic Process instead peaked at intermediate risk before declining in highest-risk, indicating that not all case-enriched pathways tracked risk uniformly. Overall, these analyses suggest pathway importance varies continuously across the predicted-risk spectrum, indicating that the models capture graded biological differences across individuals rather than average differences between cases and controls.

### 3.5. Foundation Model Biological Validation

To independently evaluate whether the biological mechanisms identified by the predictive models reflected regulatory biology, the highest- and lowest-ranked pathways from the GWAS + PRS + covs global attention model were mapped to the 76 LD-clumped AF GWAS lead variants and evaluated using the AlphaGenome foundation model across 5 GTEx tissues. These pathway scores contained no GReX-derived features, allowing for an independent assessment of tissue specificity and pathway prioritization without biasing the AlphaGenome model towards GTEx-derived information. Predicted RNA-seq regulatory effects were compared between highly and weakly attended pathways, and AlphaGenome identified significant regulatory enrichment exclusively in the two cardiac tissues comprising the primary GReX feature set. Predicted regulatory effects were significantly greater for highly attended pathways in HAA (FDR = 0.032) and HLV (FDR = 0.032), whereas no significant enrichment was observed in WB (FDR = 0.398), AC (FDR = 0.534), or AA (FDR = 0.555) (**Figure 5; Supplemental Figure S8**). These results also provide independent support for the pathway prioritization learned by the attention model. Variants mapping to highly attended pathways exhibited stronger predicted transcriptional regulatory effects than variants assigned to low-attention pathways, indicating that the model preferentially prioritized pathways enriched for regulatory activity. AlphaGenome analysis independently provides unbiased support for tissue specificity and pathway prioritization, indicating that the biological signals learned were preferentially associated with transcriptional regulatory effects in the prioritized cardiac tissues.

**Figure 5.**
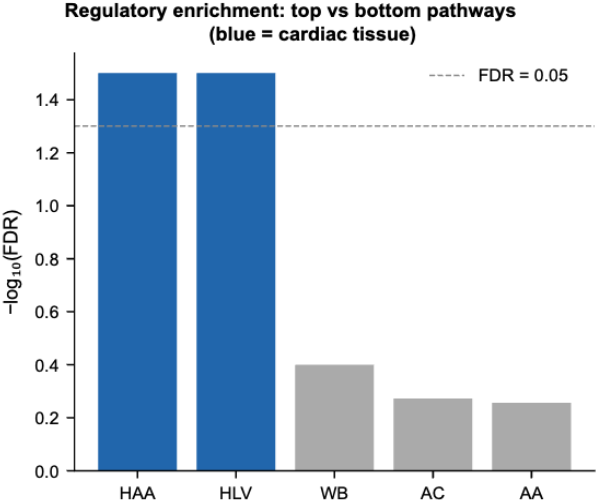
AlphaGenome transcriptional regulatory enrichment of top and bottom pathways in key tissues.

## 4. Discussion

### 4.1. Pathway-based representations improve AF prediction and interpretability

Across multiple data modalities and models, pathway-level aggregation of GWAS signals outperformed PRS (AUROC = 0.601 for PRS alone, 0.726 for PRS + covs), with the best performing models achieving AUROCs of 0.736–0.739 for the combined feature set. The similarity in performance across the linear and nonlinear models suggests that the prediction ceiling was determined largely by feature representation rather than by model complexity. Incorporating GReX from cardiac tissues did not significantly improve discrimination. Both the HAA + HLV feature set and HAA + HLV + AA sensitivity analysis performed equivalently to GWAS pathway scores alone, suggesting that pathway-level genomic features capture much of the predictive information available in this cohort. Transcriptomic information principally contributed additional biological context for improved interpretability. The HAA and HLV GReX profiles shared relatively little overlap among their highest-ranked pathways despite representing closely related cardiac tissues, demonstrating that tissue-specific transcriptomic features provide orthogonal but complementary biological information. This additional information changed how the deep learning models organized biological importance. This additional information altered attribution behavior across both deep learning architectures. In the transformer, cardiac GReX resolved an attention-sink artifact observed under GWAS + PRS alone, redistributing attention across multiple biologically coherent pathway families instead of concentrating on a single poorly interpretable pathway. In the GNN, transcriptomic augmentation reduced preferential attribution toward isolated, low-connectivity pathways (degree–GradCAM correlation: r = −0.29 to −0.03), indicating that pathway importance became substantially less dependent on graph topology. While these structural improvements were not reflected in AUROC, they suggest that transcriptomic information improves the biological fidelity of model attribution while preserving performance.

### 4.2. Population- and individual-level interpretation reveal complementary mechanisms

Interpretability analyses identified AF as a biologically heterogeneous disease involving multiple complementary processes. Across the cohort, the global attention model highlighted pathways involved in DNA repair, homologous recombination, nonsense-mediated decay, and genome maintenance, supporting growing evidence that oxidative stress and impaired DNA damage responses contribute to atrial remodeling and AF progression^18,57^. Transformer and GNN analyses converged on pathways related to cardiac electrophysiology and structural remodeling, including potassium signaling, gap junction assembly, ion transport, and sulfur compound biosynthesis^16,57,58^. The prominence of sulfur compound biosynthetic pathways is notable given increasing evidence that hydrogen sulfide regulates cardiac potassium channels and suppresses TGF-β-mediated atrial fibrosis^59–61^. Incorporating GReX from AA tissue shifted the dominant case-associated pathway toward TGF-β signaling, suggesting that sulfur metabolism and profibrotic remodeling may represent interconnected components of a broader disease process^59,62^.

These population-level findings were complemented by individual-level analyses. Transformer attention embeddings organized individuals according to pathway utilization and predicted risk rather than simple case-control status. Risk-decile analysis demonstrated that Sulfur Compound Biosynthetic Process exhibited the strongest continuous association with predicted risk (Δ = 1.65), indicating that this pathway varies progressively across patients rather than reflecting only an average case-control difference. The largest gradient under GWAS + PRS alone was significantly smaller (Δ = 0.32), suggesting that transcriptomic augmentation substantially strengthened patient-level biological stratification. The transformer and GNN highlighted complementary rather than identical aspects of disease biology, reinforcing that AF susceptibility likely arises through multiple partially overlapping molecular mechanisms rather than a single conserved pathway.

These biological observations were independently supported by AlphaGenome analysis. Variants mapping to the highest-ranked pathways from the GWAS + PRS global attention model demonstrated significantly greater predicted regulatory effects specifically within HAA and HLV, while no enrichment was observed in WB or other vascular tissues. Since this analysis did not incorporate GReX-derived features used for prediction, it provides orthogonal support that the pathways prioritized preferentially influence transcriptional regulation in disease-relevant cardiac tissues. Together, these findings suggest that transcriptomic augmentation primarily enhances biological resolution, revealing complementary population- and patient-level mechanisms while independently supporting the cardiac tissue specificity underlying those pathway associations.

### 4.3. Future Approaches and Limitations

There are several limitations in this study. This was a single-cohort study and external validation in larger, more ancestrally diverse cohorts will be important to establish the generalizability of both the pathway-level and tissue-specific findings. Second, the interpretability approaches used identify features that contribute to model predictions rather than establishing causal mechanisms. Pathway attribution varied across model architectures, indicating that some pathway-level findings may be architecture-specific and should be interpreted alongside the broader mechanisms that recurred across analyses rather than as definitive evidence for individual pathways^63^. Future work will extend these findings in several directions. Causal inference via Mendelian randomization and pathway-level mediation analyses could help determine whether the DNA repair, sulfur compound biosynthesis, and signaling pathways identified contribute directly to AF pathogenesis rather than representing correlated disease signatures. Expanding GReX to additional disease-relevant tissues may further improve biological resolution, given the complementary information observed between HAA and HLV^64^. Finally, integrating richer longitudinal clinical phenotypes, such as echocardiographic measures, with pathway-based genomic representations may improve both individualized risk prediction and mechanistic characterization of AF susceptibility^65^.

## Data Availability

All data produced in the present study are available upon reasonable request to the authors. Supplemental figures, tables, and code are available online.

https://github.com/rvenkatesh99/AFib_Pathway_Risk_Modeling

## 5. Acknowledgements

We acknowledge the Penn Medicine BioBank and Regeneron Genetics Center for providing data and thank the patient-participants who consented to participate in this research program. The PMBB is approved under IRB protocol# 813913 and supported by the Perelman School of Medicine at the University of Pennsylvania, a gift from the Smilow family, and the National Center for Advancing Translational Sciences of the NIH award #UL1TR001878.

## 6. Appendix

Supplemental figures, tables, and code are available at: https://github.com/rvenkatesh99/AFib_Pathway_Risk_Modeling

