## Supplemental Figures S1-8 for "Pathway Modeling of Genomic and Tissue-Specific Transcriptomic Architecture Identifies Personalized Mechanisms of Atrial Fibrillation Risk^†^"

#### Supplemental Figure S1: Genome-Wide Association Study Results

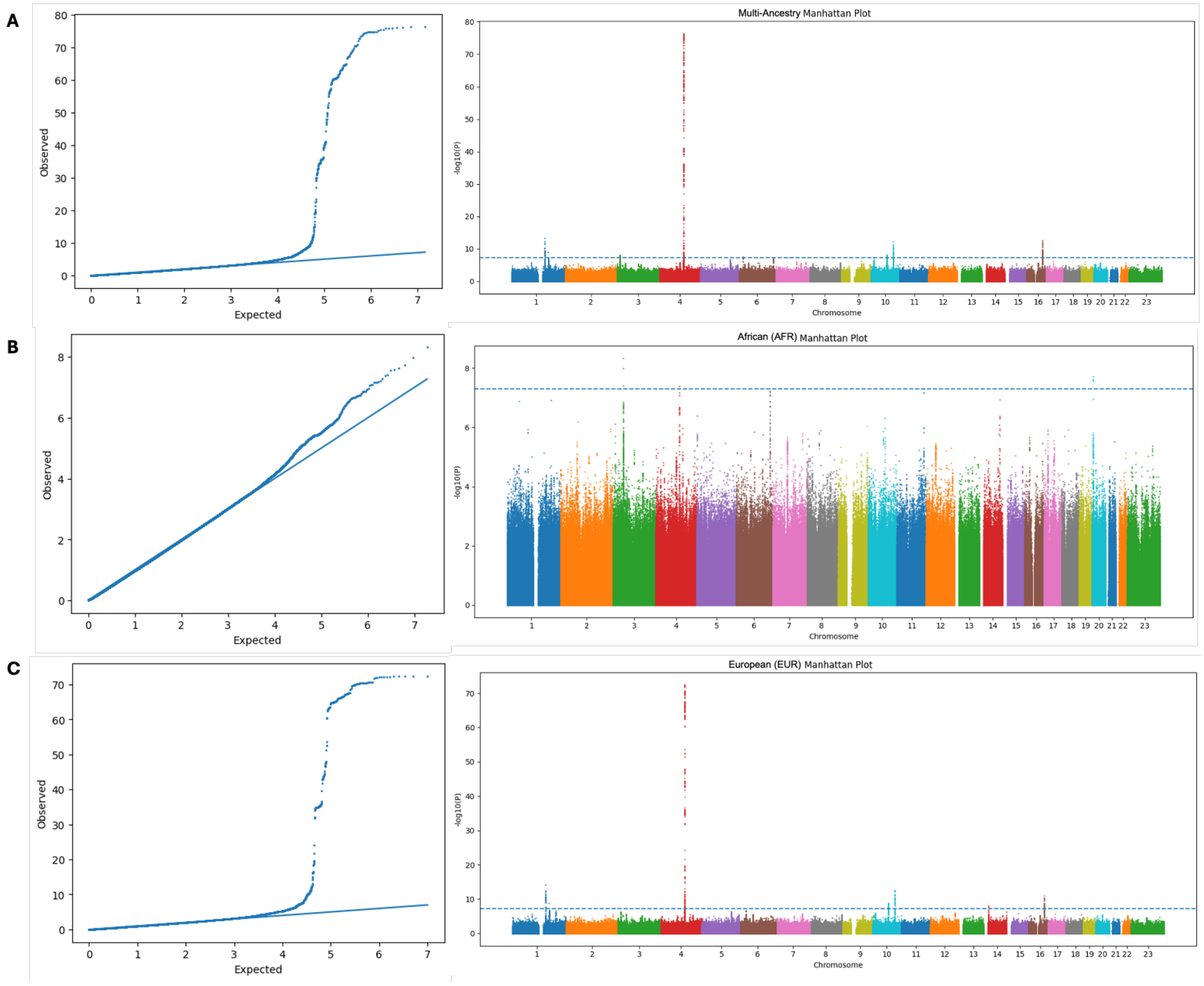

### Supplemental Figure S2: GWAS Pathway Ranking

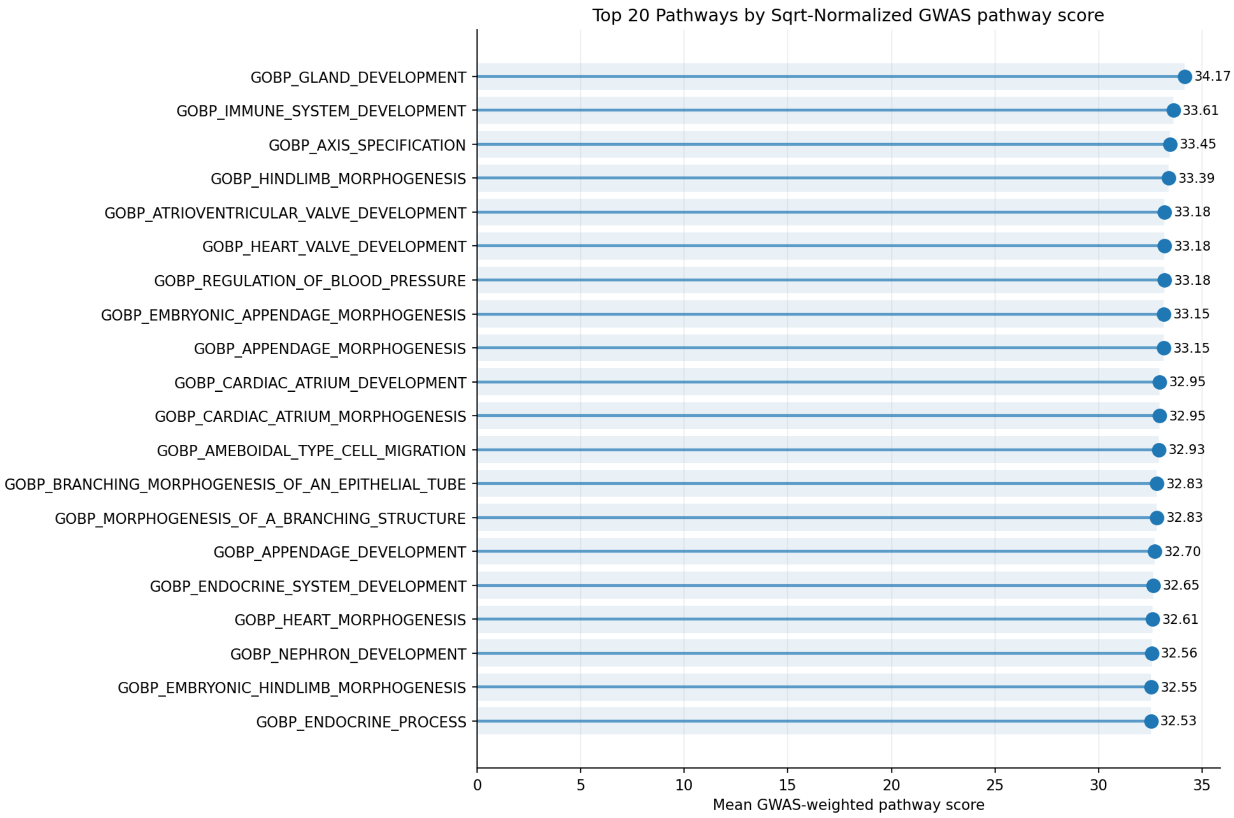


### Supplemental Figure S3: Tissue-Specific GREx Pathway Ranking

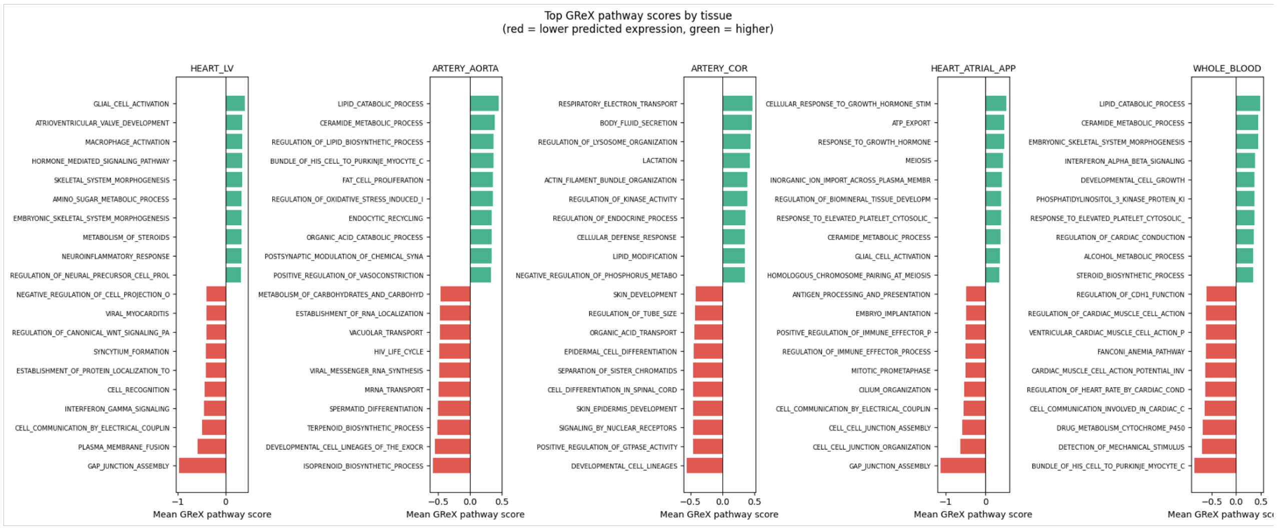

### Supplemental Figure S4: AUROC Curves

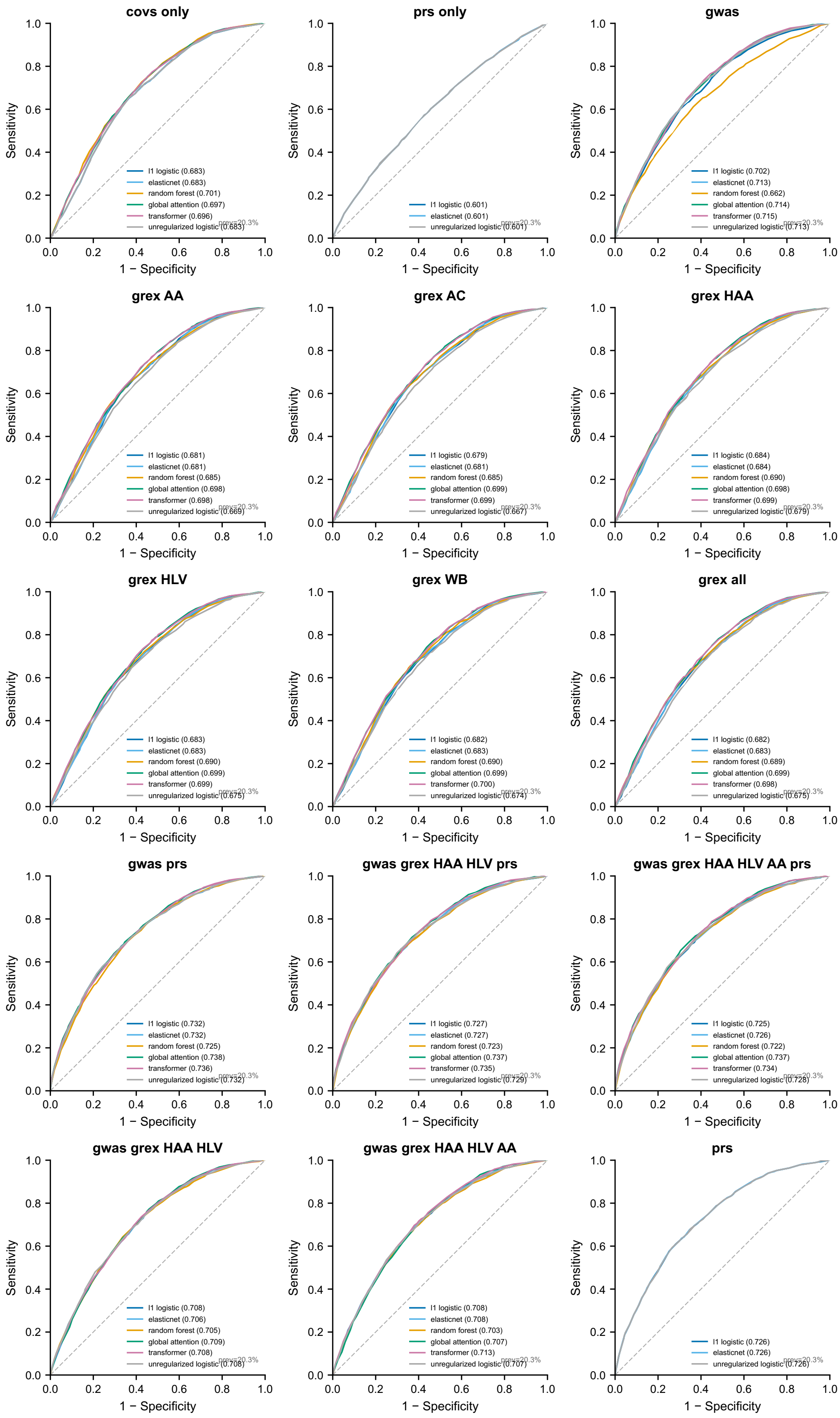

### Supplemental Figure S5: AUPRC Curves

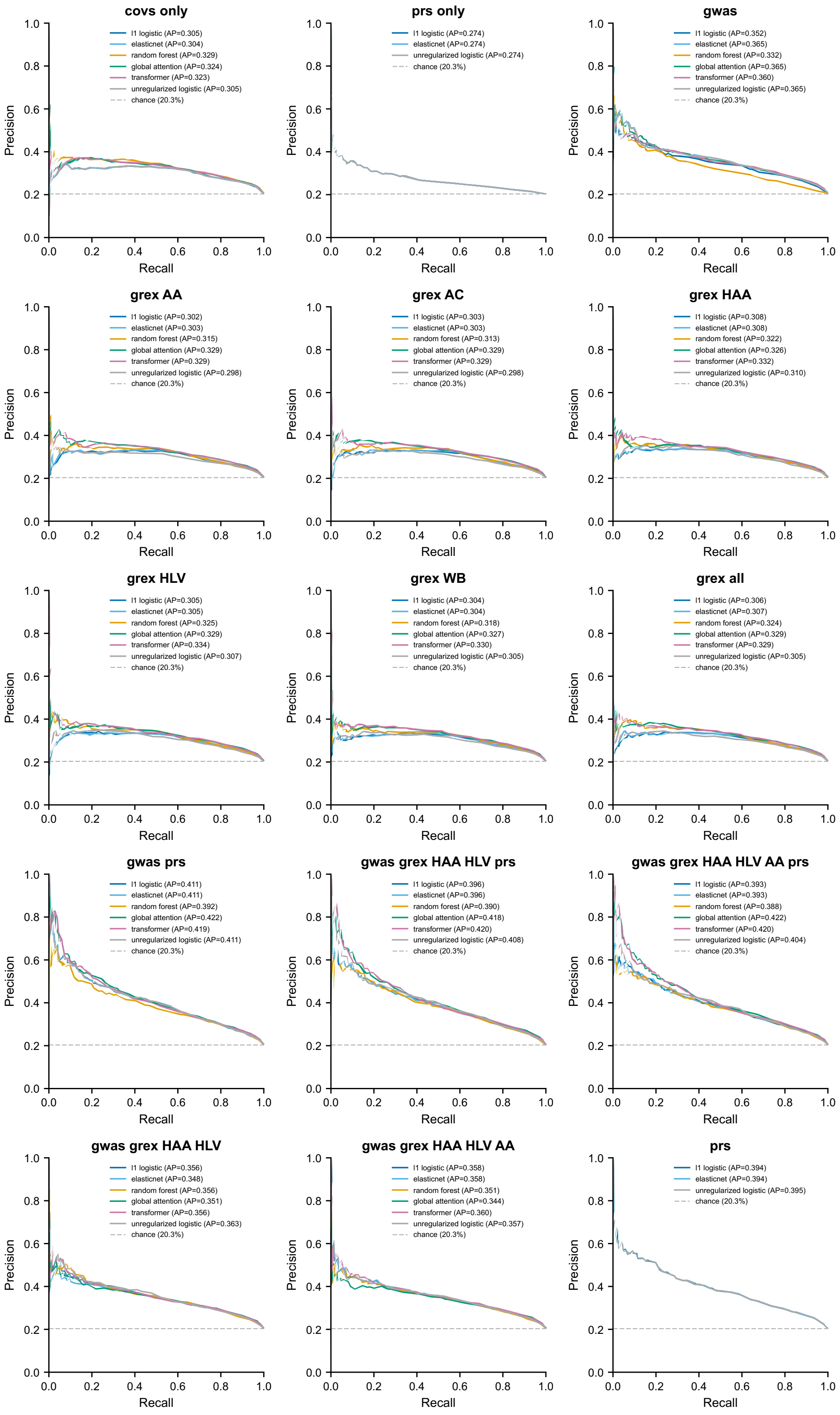

### Supplemental Figure S6: Calibration Curves

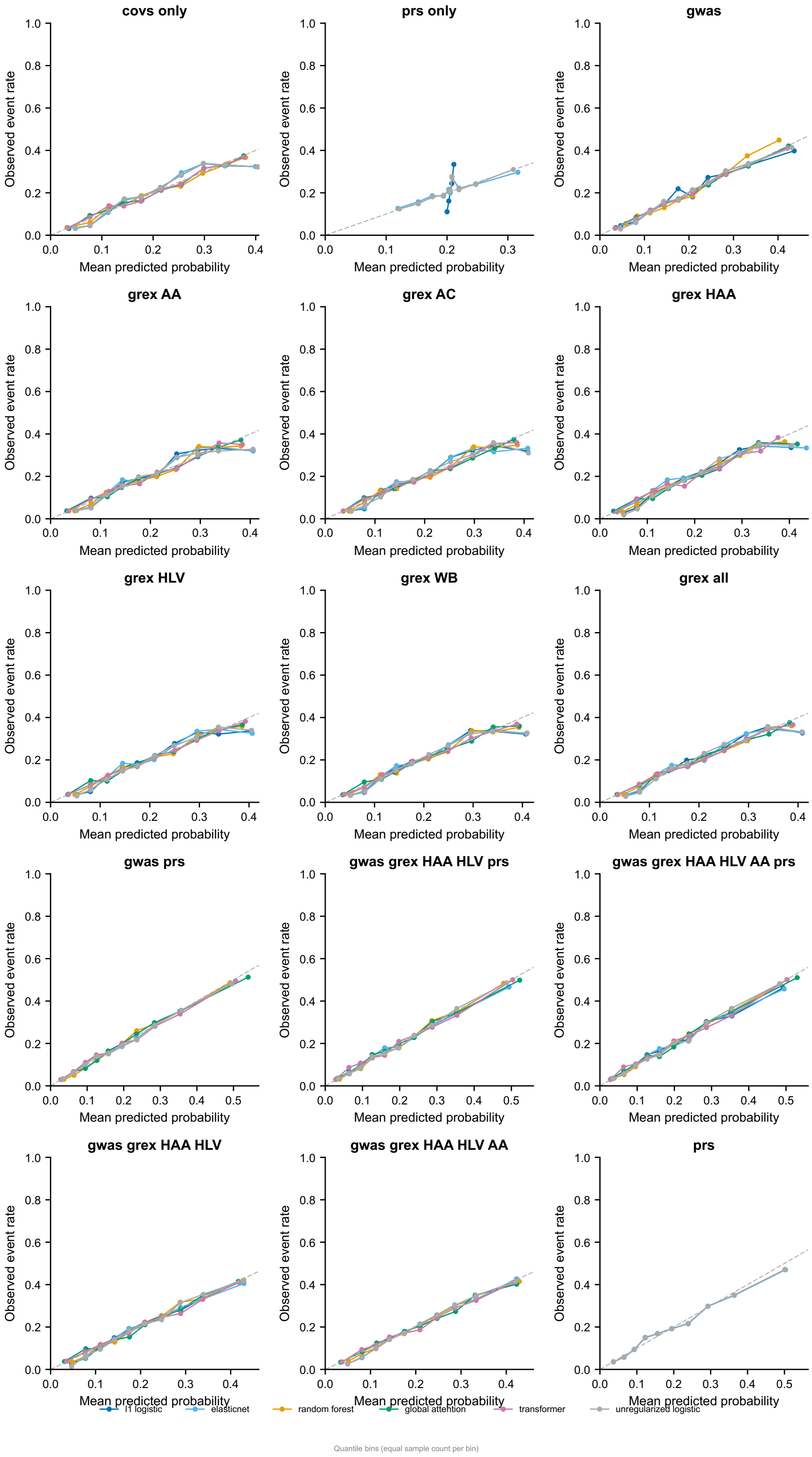

**Supplemental Figure S7: Transformer Case-Enriched Lollipop Plot for GWAS + GreX (HAA + HLV) + PRS**

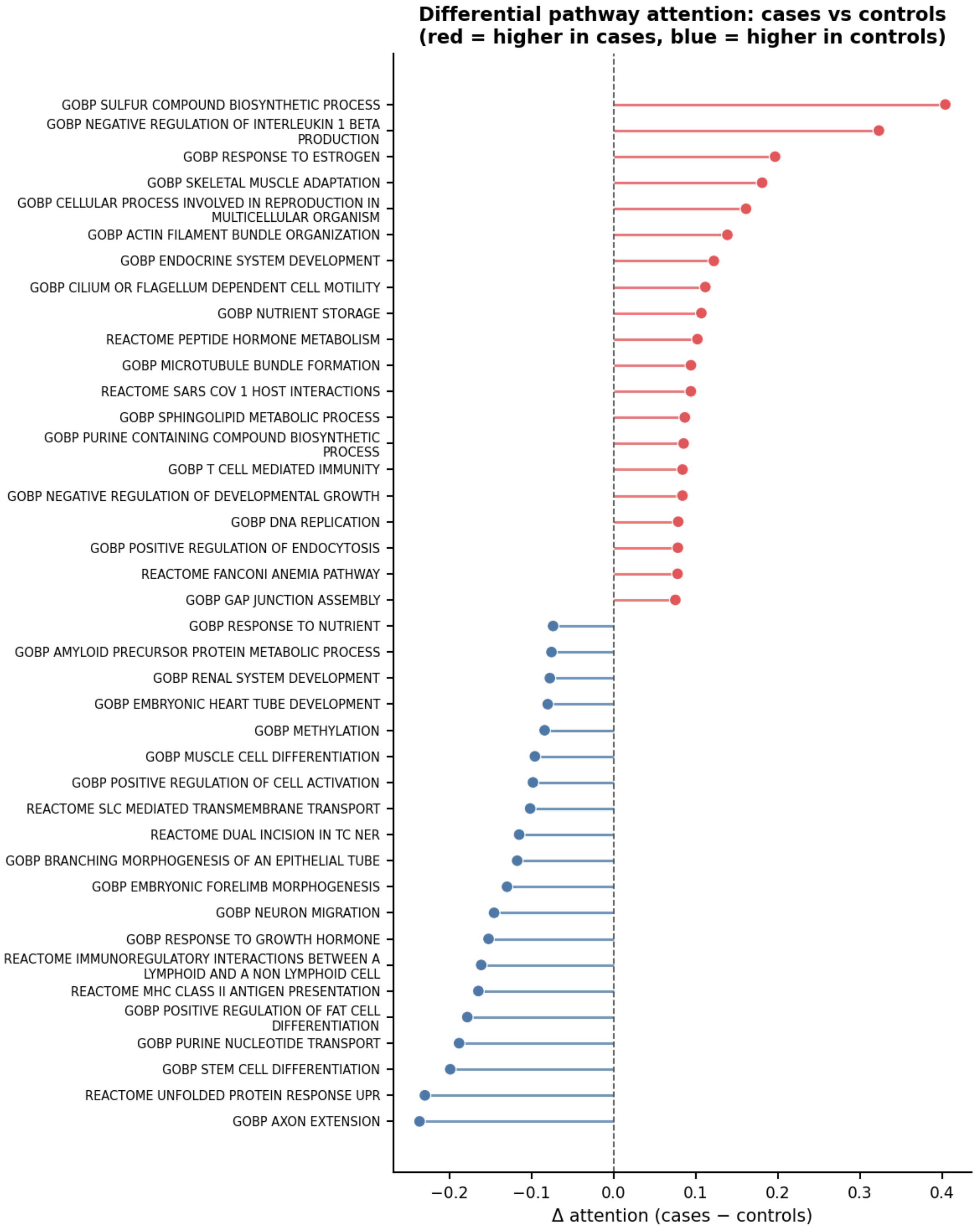

### Supplemental Figure S8: AlphaGenome Tissue-Specific Regulatory Effects

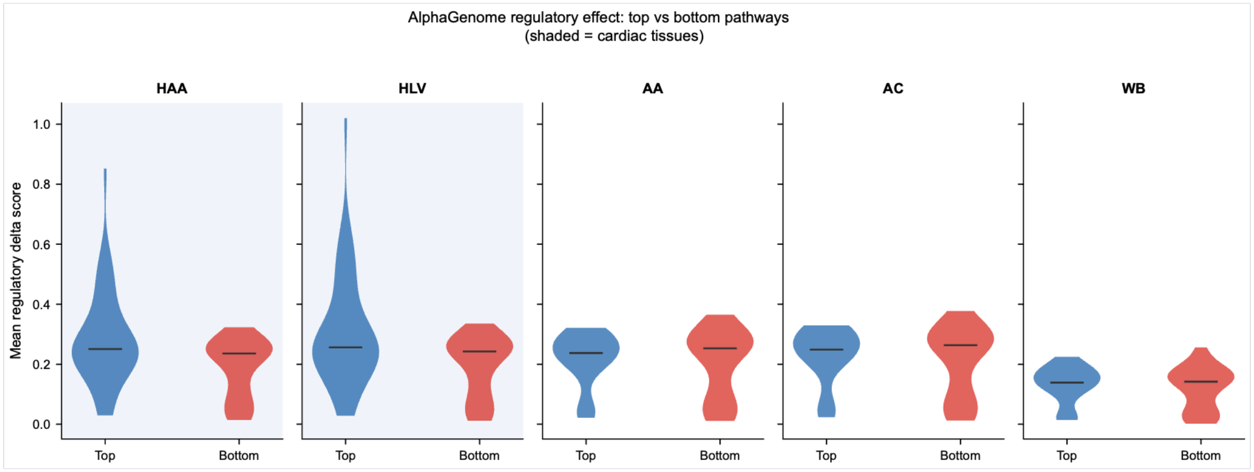
